# Research Letter: Childhood Adversity and Risk of Subsequent Head and Neck Injury: an ABCD Cohort Analysis

**DOI:** 10.64898/2025.11.28.25341166

**Authors:** D Jain, E Carlsson, NL de Souza

**Affiliations:** Department of Rehabilitation and Human Performance, Icahn School of Medicine at Mount Sinai, New York, NY; Traumatic Brain Injury and Concussion Center, University of Utah, Salt Lake City, UT

**Keywords:** pediatric, childhood adversity, brain injury, head trauma

## Abstract

**Objective:** This study examined whether baseline reports of adverse childhood experiences (ACEs) and early life adversity (ELA) were associated with subsequent head/neck injury or probable brain injury in the Adolescent Brain Cognitive Development (ABCD) Study^®^ cohort.

**Setting:** ABCD Study^®^ cohort

**Participants:** ABCD Study^®^ data release 5.0 provided baseline and partial longitudinal data through Year 4. Inclusion required at least one follow-up timepoint, data to compute the ACEs or ELA score, sex, and attention-deficit/hyperactivity disorder (ADHD) diagnosis status. The final sample included 10,853 participants for ACEs analyses and 10,850 ELA analyses.

**Design:** Prospective observational design.

**Main Measures:** Head and/or neck injuries and probable brain injuries were assessed using the caregiver-reported Ohio State University Traumatic Brain Injury Identification Method Short Form. We used previously published recommendations for the ABCD study to calculate a proxy ACEs score and a broader measure of ELA using both caregiver and youth report at baseline.

Age, gender, race/ethnicity, primary caregiver education, child’s history of ADHD, report of head/neck or probable brain injury at baseline, and broken bone injury at baseline or follow-up were covariates in logistic regression models examining the association between risk of head/neck injury or probable brain injury with ACEs or ELA.

**Results:** Exposure to more adversity (ACES: Odds Ratio [95% Confidence Interval] OR[95%CI]=0.04[0.001,0.08]; ELA: OR[95%CI]=0.05[0.01,0.08]) was associated with higher odds of sustaining a head or neck injury at any of the follow-up time points but not of probable brain injury (ACES: OR[95%CI]=0.002[−0.08,0.08]; ELA: OR[95%CI]=0.01[−0.06,0.07]).

**Conclusion:** Limited public knowledge of and reliance on caregiver report of head/neck injury increases misclassification bias and underscores the need for improved education and measurement strategies. We observed a stronger association between ELA and head/neck injury, potentially reflecting differences in disclosure and awareness rather than due to types of exposure.

## Introduction

Childhood and adolescence are periods of rapid social, emotional, and cognitive growth that can shape long-term health and development. Early life adversity (ELA) can negatively alter developmental trajectories and includes a broad range of harmful experiences inside (e.g., physical, emotional, or sexual abuse, neglect, parental substance misuse) and outside the household (e.g., exposure to disaster and community violence, family separation, lack of safety or basic resources).^1^ Adverse childhood experiences (ACEs) fall within this umbrella term and specifically encompass within-household experiences. Both ACEs and ELA have been associated with increased risk of mental and physical health concerns, including greater adult morbidity and mortality,^2^ cognitive deficits, and frequent mental distress.^3^ A recent meta-analysis of 206 studies across 22 countries reported 60% of adults endorsed at least one ACE, and 16% endorsed four or more before the age of 18,^4^ highlighting the significant public health concern.

Traumatic brain injury (TBI) affects an estimated 5.4% of American children, with one in three experiencing persistent somatic, cognitive, sleep, and psychological symptoms.^5-8^ These persistent symptoms significantly reduce quality of life and can disrupt typical social and developmental activities like attending school.^9^

Two studies have linked ACEs to increased TBI risk. In the Adolescent Brain Cognitive Development (ABCD) Study cohort, analyses of baseline data found that more ACEs were associated with greater odds of both a lifetime head or neck injury and TBI among children aged 9-10 years.^10^ Children with four or more ACEs had 70-140% increased odds of head or neck injury, compared to those with none.^10^ The National Survey of Children’s Health similarly found that adolescents who reported four or more ACEs had a 347% increased risk of TBI.^11^

Despite these findings, limited research has investigated how ACEs and ELAs predict TBI risk longitudinally among children. The current study uses the ABCD Study^®^ cohort to examine whether baseline ACEs and ELA are associated with mild head or neck injury at follow-up. We hypothesized that greater early life adversity would increase subsequent TBI risk.

## Methods

### Design

We used data release 5.0 of the ABCD Study^®^, which includes baseline data and partial longitudinal data through the Year 4 follow-up visit. The present study analyzed repeated caregiver- and youth-reported measures collected over this period. This secondary data analysis was determined to be exempt by the Institutional Review Board at the Icahn School of Medicine at Mount Sinai.

### Sample

The analytic sample included participants who had data from at least one follow-up visit. Participants were excluded if they were missing questionnaire responses needed to compute the Adverse Childhood Experiences (ACEs) proxy score (n=877) or the Early Life Adversity (ELA) score (n=880). Additionally, three participants were removed for not endorsing female or male sex, and one participant was removed due to missing attention-deficit/hyperactivity disorder (ADHD) diagnosis at baseline. The final sample included 10,853 participants for ACEs analyses and 10,850 participants for ELA analyses.

### Measures

#### Outcome variables

Head and/or neck (hereafter, head/neck) injuries and probable brain injuries were assessed using the caregiver-reported Ohio State University Traumatic Brain Injury Identification Method Short Form (OSU-TBI-ID). Caregivers indicated whether the child had experienced any of the following four injury scenarios: a head/neck injury (1) requiring hospital or emergency department treatment; (2) resulting from a motor vehicle or other moving-vehicle crash; (3) from a fall, being hit, or sports participation; and (4) from a fight or being shaken violently. For each endorsed scenario, caregivers reported the presence and duration of loss of consciousness, as well as whether the child was dazed or experienced a memory gap following the injury. Injuries involving loss of consciousness, being dazed, or a memory gap were classified as probable brain injuries, whereas injuries without these post-injury symptoms were classified as head/neck injuries. At baseline, caregivers reported lifetime history, and at subsequent follow-up visits they reported any injuries that occurred since the previous visit. Final outcomes were coded as binary variables: 0 indicated no head/neck or probable brain injury reported across all follow-up timepoints, and 1 indicated the occurrence of any head/neck or probable brain injury.

#### Exposure Variables

We used Breslin et al., 2025 best practices for measuring adversity to calculate both a proxy ACEs score and a broader measure of ELA using caregiver and youth reports at baseline.^1^ The ELA score includes exposure, lack of safety, lack of economic resources, and separation from caregiver(s) in addition to the traditional ACEs domains.

#### Covariates

We included child age, gender, race/ethnicity, and primary caregiver education, child’s history of ADHD, and report of head/neck or probable brain injury at baseline, as these characteristics have been associated with head/neck or probable brain injury. We also included whether a broken bone injury was reported at baseline or follow-up to ensure that any relationships were not reflective of overall injury incidence, but specific to injuries to the head or neck.

#### Analyses

Analyses were conducted using R version 4.4.2. Multiple logistic regressions were used to estimate the odds of 1) head/neck injury, and 2) probable brain injury from the ACEs proxy score or ELA score while adjusting for the covariates. Odds ratios (OR) and 95% confidence intervals (95%CI) are reported for each model.

## Results

Demographic characteristics of the analytic sample can be found in Table 1.

**Table 1.**
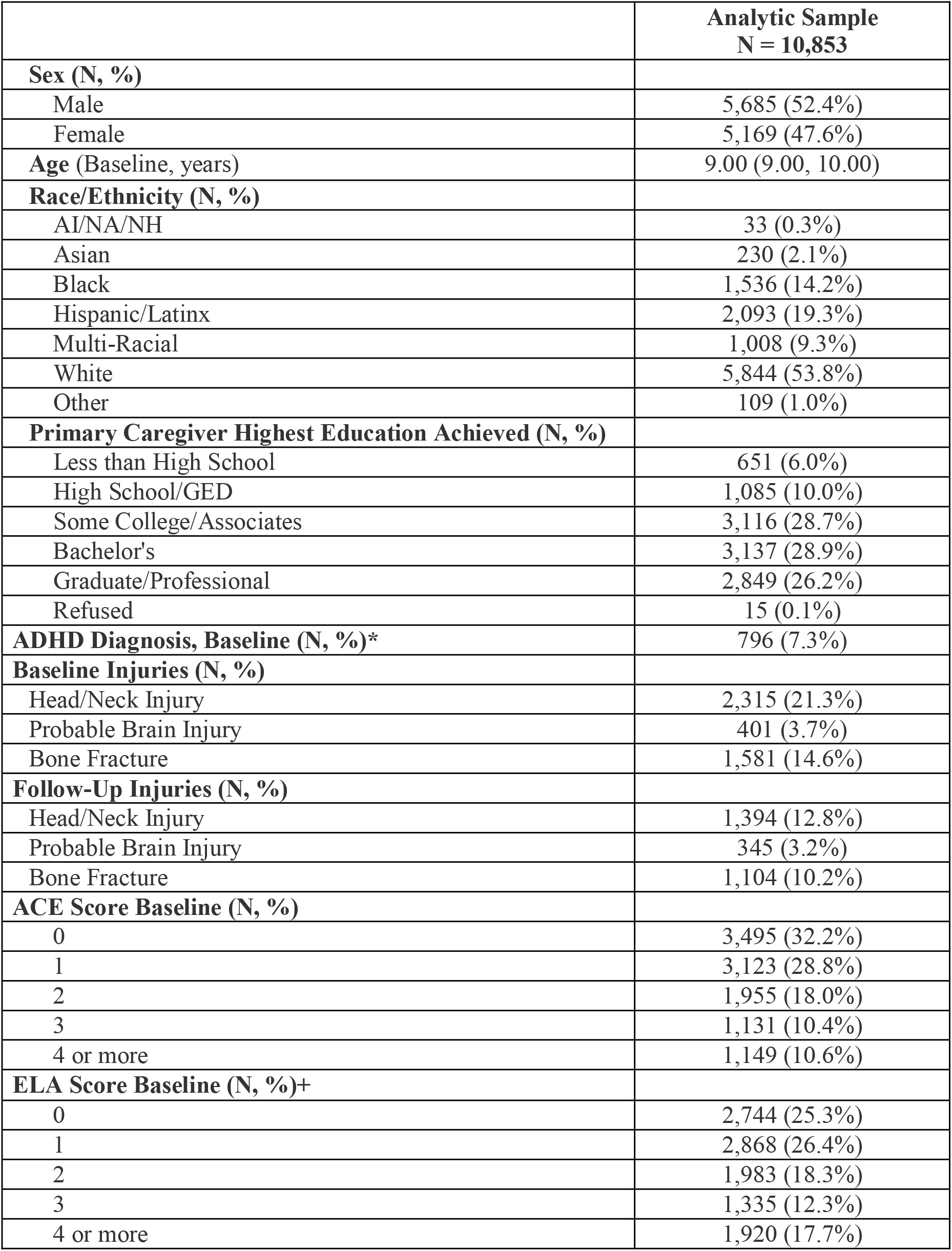
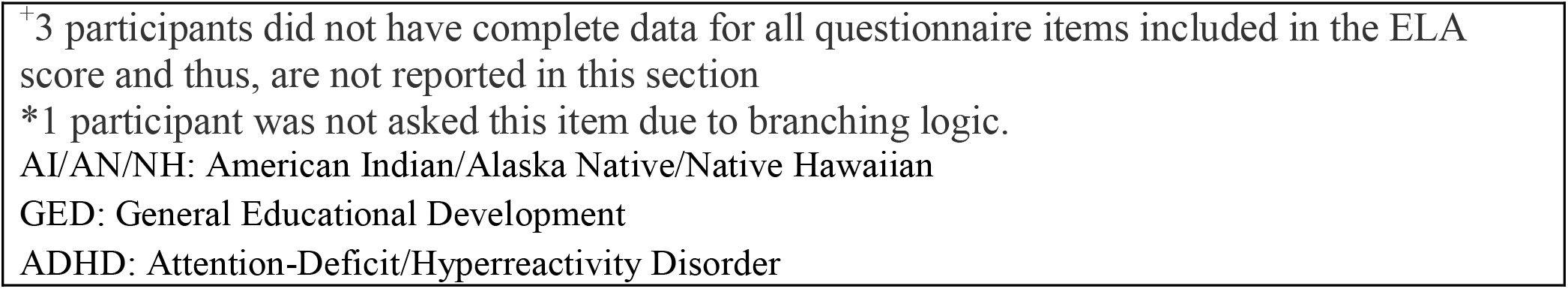
Demographic + variable characteristics for the full sample. Summaries are presented as either number (N) and proportion (%), or median and lower and upper quartiles (Q1, Q3).

|  | **Analytic Sample**<br>**N = 10,853** |
| --- | --- |
| **Sex (N, %)** |  |
| Male | 5,685 (52.4%) |
| Female | 5,169 (47.6%) |
| **Age (Baseline, years)** | 9.00 (9.00, 10.00) |
| **Race/Ethnicity (N, %)** |  |
| AI/NA/NH | 33 (0.3%) |
| Asian | 230 (2.1%) |
| Black | 1,536 (14.2%) |
| Hispanic/Latinx | 2,093 (19.3%) |
| Multi-Racial | 1,008 (9.3%) |
| White | 5,844 (53.8%) |
| Other | 109 (1.0%) |
| **Primary Caregiver Highest Education Achieved (N, %)** |  |
| Less than High School | 651 (6.0%) |
| High School/GED | 1,085 (10.0%) |
| Some College/Associates | 3,116 (28.7%) |
| Bachelor's | 3,137 (28.9%) |
| Graduate/Professional | 2,849 (26.2%) |
| Refused | 15 (0.1%) |
| **ADHD Diagnosis, Baseline (N, %)\*** | 796 (7.3%) |
| **Baseline Injuries (N, %)** |  |
| Head/Neck Injury | 2,315 (21.3%) |
| Probable Brain Injury | 401 (3.7%) |
| Bone Fracture | 1,581 (14.6%) |
| **Follow-Up Injuries (N, %)** |  |
| Head/Neck Injury | 1,394 (12.8%) |
| Probable Brain Injury | 345 (3.2%) |
| Bone Fracture | 1,104 (10.2%) |
| **ACE Score Baseline (N, %)** |  |
| 0 | 3,495 (32.2%) |
| 1 | 3,123 (28.8%) |
| 2 | 1,955 (18.0%) |
| 3 | 1,131 (10.4%) |
| 4 or more | 1,149 (10.6%) |
| **ELA Score Baseline (N, %)+** |  |
| 0 | 2,744 (25.3%) |
| 1 | 2,868 (26.4%) |
| 2 | 1,983 (18.3%) |
| 3 | 1,335 (12.3%) |
| 4 or more | 1,920 (17.7%) |

<sup>+</sup>3 participants did not have complete data for all questionnaire items included in the ELA score and thus, are not reported in this section
\*1 participant was not asked this item due to branching logic.
AI/AN/NH: American Indian/Alaska Native/Native Hawaiian
GED: General Educational Development
ADHD: Attention-Deficit/Hyperactivity Disorder

After adjusting for covariates, exposure to more adversity (ACES: OR[95% CI] = OR[95%CI]=0.04[0.001,0.08]; ELA: OR[95%CI]=0.05[0.01,0.08]) was associated with higher odds of sustaining a head or neck injury at any of the follow-up time points but not of probable brain injury (ACES: OR[95%CI]=0.002[−0.08,0.08]; ELA: OR[95%CI]=0.01[−0.06,0.07]). ORs and 95% CIs for all variables can be found in Tables 2 and 3.

**Table 2.**
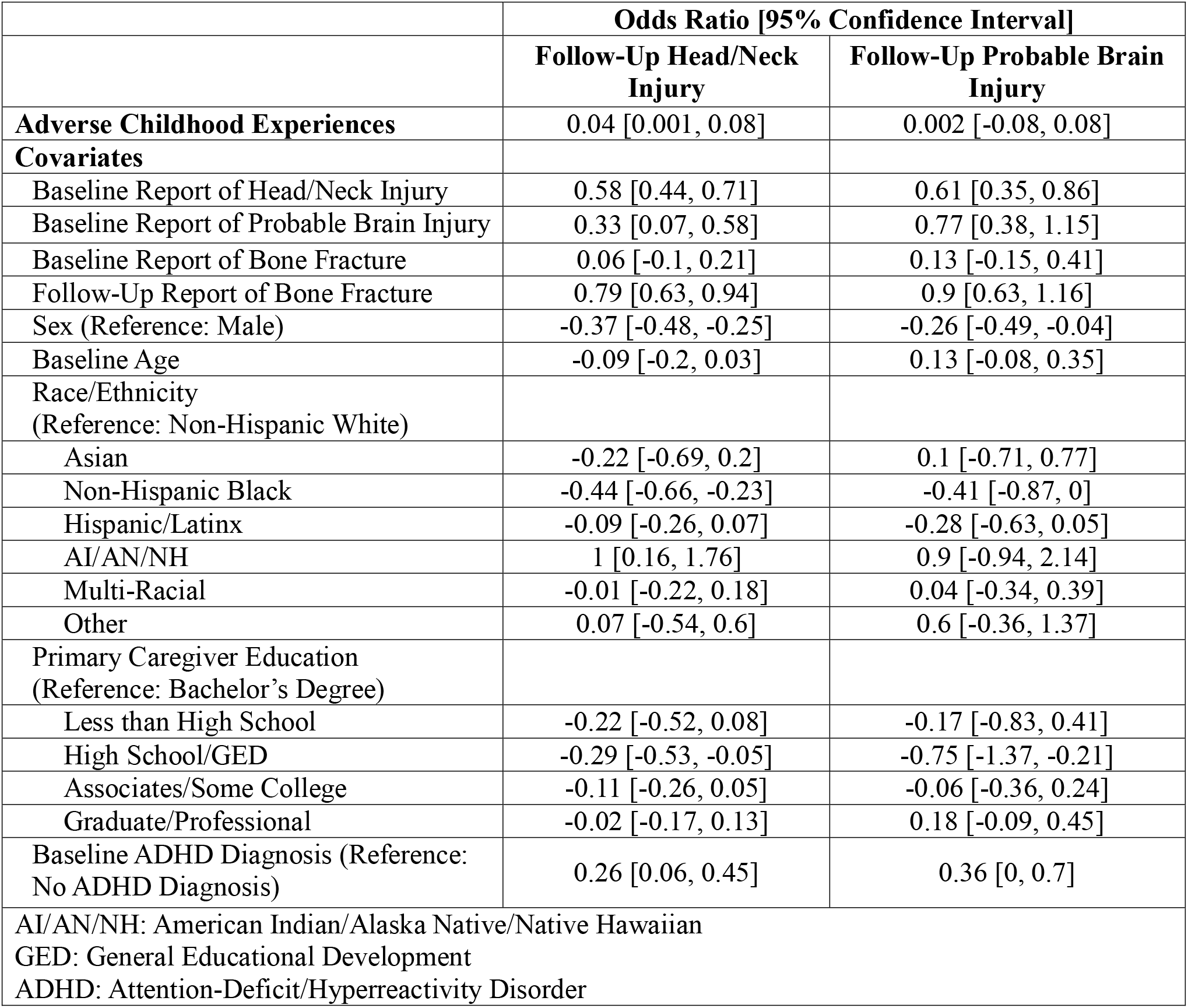
Odds Ratios for models with total adverse childhood experiences score.

|  | **Odds Ratio [95% Confidence Interval]** |  |
| --- | --- | --- |
|  | **Follow-Up Head/Neck Injury** | **Follow-Up Probable Brain Injury** |
| **Adverse Childhood Experiences** | 0.04 [0.001, 0.08] | 0.002 [-0.08, 0.08] |
| **Covariates** |  |  |
| Baseline Report of Head/Neck Injury | 0.58 [0.44, 0.71] | 0.61 [0.35, 0.86] |
| Baseline Report of Probable Brain Injury | 0.33 [0.07, 0.58] | 0.77 [0.38, 1.15] |
| Baseline Report of Bone Fracture | 0.06 [-0.1, 0.21] | 0.13 [-0.15, 0.41] |
| Follow-Up Report of Bone Fracture | 0.79 [0.63, 0.94] | 0.9 [0.63, 1.16] |
| Sex (Reference: Male) | -0.37 [-0.48, -0.25] | -0.26 [-0.49, -0.04] |
| Baseline Age | -0.09 [-0.2, 0.03] | 0.13 [-0.08, 0.35] |
| Race/Ethnicity<br>(Reference: Non-Hispanic White) |  |  |
| Asian | -0.22 [-0.69, 0.2] | 0.1 [-0.71, 0.77] |
| Non-Hispanic Black | -0.44 [-0.66, -0.23] | -0.41 [-0.87, 0] |
| Hispanic/Latinx | -0.09 [-0.26, 0.07] | -0.28 [-0.63, 0.05] |
| AI/AN/NH | 1 [0.16, 1.76] | 0.9 [-0.94, 2.14] |
| Multi-Racial | -0.01 [-0.22, 0.18] | 0.04 [-0.34, 0.39] |
| Other | 0.07 [-0.54, 0.6] | 0.6 [-0.36, 1.37] |
| Primary Caregiver Education<br>(Reference: Bachelor's Degree) |  |  |
| Less than High School | -0.22 [-0.52, 0.08] | -0.17 [-0.83, 0.41] |
| High School/GED | -0.29 [-0.53, -0.05] | -0.75 [-1.37, -0.21] |
| Associates/Some College | -0.11 [-0.26, 0.05] | -0.06 [-0.36, 0.24] |
| Graduate/Professional | -0.02 [-0.17, 0.13] | 0.18 [-0.09, 0.45] |
| Baseline ADHD Diagnosis (Reference:<br>No ADHD Diagnosis) | 0.26 [0.06, 0.45] | 0.36 [0, 0.7] |
| AI/AN/NH: American Indian/Alaska Native/Native Hawaiian<br>GED: General Educational Development<br>ADHD: Attention-Deficit/Hyperactivity Disorder |  |  |

**Table 3.**
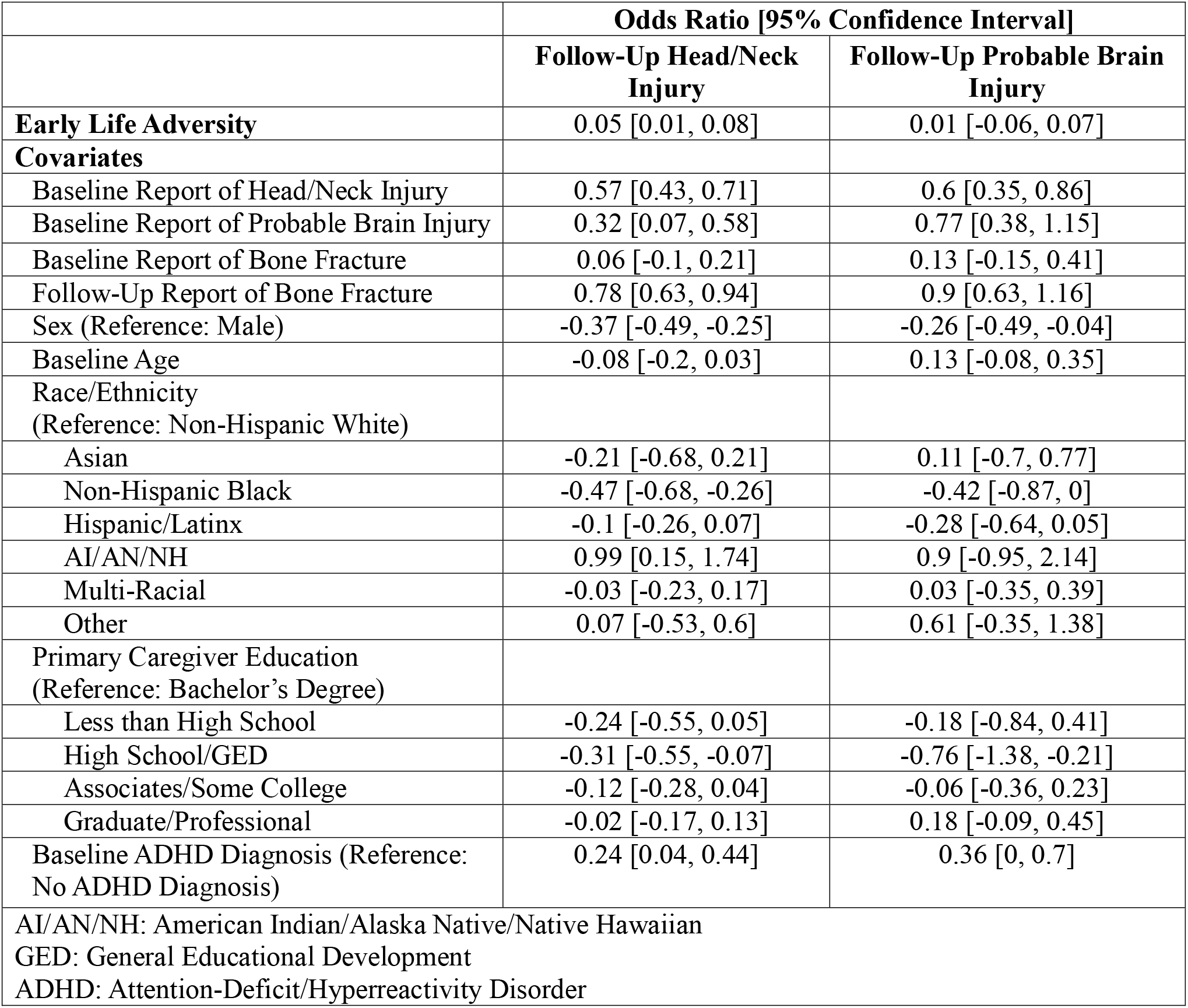
Odds ratios for models with total early life adversity score.

|  | **Odds Ratio [95% Confidence Interval]** |  |
| --- | --- | --- |
|  | **Follow-Up Head/Neck Injury** | **Follow-Up Probable Brain Injury** |
| **Early Life Adversity** | 0.05 [0.01, 0.08] | 0.01 [-0.06, 0.07] |
| **Covariates** |  |  |
| Baseline Report of Head/Neck Injury | 0.57 [0.43, 0.71] | 0.6 [0.35, 0.86] |
| Baseline Report of Probable Brain Injury | 0.32 [0.07, 0.58] | 0.77 [0.38, 1.15] |
| Baseline Report of Bone Fracture | 0.06 [-0.1, 0.21] | 0.13 [-0.15, 0.41] |
| Follow-Up Report of Bone Fracture | 0.78 [0.63, 0.94] | 0.9 [0.63, 1.16] |
| Sex (Reference: Male) | -0.37 [-0.49, -0.25] | -0.26 [-0.49, -0.04] |
| Baseline Age | -0.08 [-0.2, 0.03] | 0.13 [-0.08, 0.35] |
| Race/Ethnicity<br>(Reference: Non-Hispanic White) |  |  |
| Asian | -0.21 [-0.68, 0.21] | 0.11 [-0.7, 0.77] |
| Non-Hispanic Black | -0.47 [-0.68, -0.26] | -0.42 [-0.87, 0] |
| Hispanic/Latinx | -0.1 [-0.26, 0.07] | -0.28 [-0.64, 0.05] |
| AI/AN/NH | 0.99 [0.15, 1.74] | 0.9 [-0.95, 2.14] |
| Multi-Racial | -0.03 [-0.23, 0.17] | 0.03 [-0.35, 0.39] |
| Other | 0.07 [-0.53, 0.6] | 0.61 [-0.35, 1.38] |
| Primary Caregiver Education<br>(Reference: Bachelor's Degree) |  |  |
| Less than High School | -0.24 [-0.55, 0.05] | -0.18 [-0.84, 0.41] |
| High School/GED | -0.31 [-0.55, -0.07] | -0.76 [-1.38, -0.21] |
| Associates/Some College | -0.12 [-0.28, 0.04] | -0.06 [-0.36, 0.23] |
| Graduate/Professional | -0.02 [-0.17, 0.13] | 0.18 [-0.09, 0.45] |
| Baseline ADHD Diagnosis (Reference:<br>No ADHD Diagnosis) | 0.24 [0.04, 0.44] | 0.36 [0, 0.7] |
| AI/AN/NH: American Indian/Alaska Native/Native Hawaiian<br>GED: General Educational Development<br>ADHD: Attention-Deficit/Hyperactivity Disorder |  |  |

## Discussion

Prior research demonstrates that both children and parents have substantial gaps in knowledge about TBI and concussion, despite increased public awareness efforts. A scoping review found that children under the age of 14, which represents most of the current sample, frequently lack understanding of concussion signs, symptoms, and reporting requirements, compared to older children. One prior study found modest concussion knowledge among parents and students, with parents averaging 70.6% correct and students 64.9%, while TBI knowledge accuracy was even lower (60% and 60.8%, respectively), with no significant group differences for TBI knowledge.^12^ These studies have important implications for our findings as head/neck injury and probable TBI classification relies solely on caregiver report in the ABCD study. If children do not recognize or communicate symptoms such as loss of or alterations in consciousness, or caregivers do not recognize the importance of such symptoms, these injuries may go unreported. Thus, the associations observed between childhood adversity and head/neck injury but not probable brain injury may partly reflect under-recognized or unreported injuries rather than a true absence of brain injury. Limited public knowledge surrounding brain injury, paired with reliance on caregiver knowledge, increases the likelihood of misclassification bias and underscores the need for improved education and more rigorous measurement strategies.

We also found that the odds ratio for ELA was slightly higher than that of ACEs in predicting subsequent head/neck injury. A study of 522 parent-teen dyads found substantial discordance between parent and teen reports of adversity, with disagreement ranging from 2.9% to 21.2%. Agreement was highest for household challenges such as parental divorce (κ = 0.69, substantial agreement), but markedly lower for abuse, neglect, and violence-related ACEs like emotional (κ = 0.27, fair agreement) and physical abuse (κ = 0.18, slight agreement).^13^ These low levels of agreement are similar to a prior examination of the ABCD study cohort finding discordance between caregiver and youth reports of social victimization.^14^ It is possible that underreporting of sensitive ACEs, especially those involving abuse, violence, or caregiver perpetration, may have attenuated associations between cumulative ACEs and injury risk. Conversely, measures of ELA, such as neighborhood violence, community safety, and discrimination, may have been easier to disclose, leading to higher endorsement rates and stronger observed associations with risk of head/neck injury. Thus, the stronger association we observed between ELA and head/neck injury, compared to ACEs, may reflect differences in disclosure and awareness rather than differences in underlying exposure.

### Limitations

The quantification of both ACEs and ELAs was determined via both caregiver and child reported measures. Although we employed a measurement method that was based on a review of all examinations of childhood adversity within the ABCD cohort^1^, we were unable to adjust for the likely discordance that exists between caregiver and child report for many measures of adversity.^14^ Other measures of childhood adversity (e.g., adversity across neighborhood or community contexts) are captured for a subset of ABCD participants via the Social Development Substudy (ABCD-SD). Because these measures were only collected in a subset of participants after the baseline or initial study visit, we were unable to incorporate them into our analyses and acknowledge that these additional measures of adversity may have changed the relationship between overall childhood adversity and risk of head/neck or probable brain injury. Additionally, we did not incorporate any potential protective factors (e.g., resilience, predictable routines etc.) into analyses. These may be important to investigate as they may moderate the relationship between childhood adversity and risk of head/neck or probable brain injury. As mentioned previously, the OSU-TBI-ID being solely a caregiver reported measure may underestimate the prevalence of these injuries. Future work may attempt to incorporate child report as well as medical records or athletic trainer or coach reports to better estimate the prevalence of head/neck and probable brain injury.

## Conclusion

Childhood adversity was associated with higher odds of later head and neck injury but not probable brain injury. These findings underscore the importance of improving public knowledge of TBI symptoms and strengthening reporting of both injury and adversity to reduce misclassification. Future research that uses objective injury markers and more detailed adversity assessments will clarify how childhood adversity relates to injury risk.

## Data Availability

The Adolescent Brain Cognitive DevelopmentSM (ABCD) Study (https://abcdstudy.org) are held in the NIMH Data Archive (NDA).

## Acknowledgements

Data used in the preparation of this article were obtained from the Adolescent Brain Cognitive Development^SM^ (ABCD) Study (https://abcdstudy.org), held in the NIMH Data Archive (NDA). This is a multisite, longitudinal study designed to recruit more than 10,000 children age 9-10 and follow them over 10 years into early adulthood. The ABCD Study® is supported by the National Institutes of Health and additional federal partners under award numbers U01DA041048, U01DA050989, U01DA051016, U01DA041022, U01DA051018, U01DA051037, U01DA050987, U01DA041174, U01DA041106, U01DA041117, U01DA041028, U01DA041134, U01DA050988, U01DA051039, U01DA041156, U01DA041025, U01DA041120, U01DA051038, U01DA041148, U01DA041093, U01DA041089, U24DA041123, U24DA041147. A full list of supporters is available at https://abcdstudy.org/federal-partners.html. A listing of participating sites and a complete listing of the study investigators can be found at https://abcdstudy.org/consortium_members/. ABCD consortium investigators designed and implemented the study and/or provided data but did not necessarily participate in the analysis or writing of this report. This manuscript reflects the views of the authors and may not reflect the opinions or views of the NIH or ABCD consortium investigators.

## Notes

**Conflicts of Interest:** The authors declare no conflicts of interest.

### Competing Interest Statement

The authors have declared no competing interest.

### Author Declarations

Data used in the preparation of this article were obtained from the Adolescent Brain Cognitive DevelopmentSM (ABCD) Study (https://abcdstudy.org), held in the NIMH Data Archive (NDA). This is a multisite, longitudinal study designed to recruit more than 10,000 children age 9-10 and follow them over 10 years into early adulthood.

